# Delta-like ligand 3 expression and functional imaging in gastroenteropancreatic neuroendocrine neoplasms

**DOI:** 10.1101/2025.06.24.25330227

**Authors:** Rohit Thummalapalli, Salomon Tendler, Joanne F. Chou, Zeynep C. Tarcan, Courtney Porfido, Jonathan Willner, Irina Linkov, Umesh Bhanot, Alissa J. Cooper, Jierui Xu, James J. Harding, Natasha Rekhtman, Laura H. Tang, Charles M. Rudin, Yelena Y. Janjigian, Heiko Schöder, John T. Porier, Jinru Shia, Olca Basturk, Diane Reidy-Lagunes, Marinela Capanu, Jason S. Lewis, Lisa Bodei, Mark P. Dunphy, Nitya Raj

## Abstract

**Purpose:** Delta-like ligand 3 (DLL3) is an emerging target across neuroendocrine cancers, but remains underexplored in gastroenteropancreatic neuroendocrine neoplasms (GEP NENs) including poorly differentiated neuroendocrine carcinomas (GEP NECs) and well differentiated neuroendocrine tumors (NETs). We aimed to define the landscape of DLL3 expression and feasibility of DLL3-targeted imaging in this population.

**Patients and Methods:** We completed DLL3 immunohistochemistry (IHC) on 360 tumor samples from patients with GEP NENs, analyzing associations between DLL3 IHC positivity and clinicopathologic features and outcomes. [^89^Zr]Zr-DFO-SC16.56 DLL3 immunoPET-CT imaging was performed in six patients with DLL3 IHC-positive advanced GEP NENs as part of a phase II clinical trial.

**Results:** Among GEP NECs, DLL3 expression was identified in 53/75 (71%) samples, was enriched for small cell histology, and did not demonstrate prognostic significance. Among well differentiated pancreatic NETs (PanNETs), DLL3 expression was identified in 22/51 (43%) grade 3 (G3) tumors, with univariate analysis revealing increased mortality risk among patients with DLL3-positive advanced G3 PanNETs (hazard ratio 3.27, 95% confidence interval 1.09-9.78). Between May 28, 2024 and February 10, 2025, six patients with DLL3 IHC-positive GEP NENs were enrolled onto the imaging protocol. [^89^Zr]Zr-DFO-SC16.56 immunoPET-CT imaging delineated DLL3-avid tumor lesions in five of six patients (two of two GEP NECs, three of four G3 PanNETs). Tumor-specific uptake of [^89^Zr]Zr-DFO-SC16.56 varied between patients, with maximum standard uptake values ranging from 7.4-36.7, with four of six cases demonstrating DLL3 avidity in ≥ 50% of tumor lesions.

**Conclusion:** DLL3 is expressed on a majority of GEP NECs and on a subset of high grade PanNETs marked by poor outcomes. Functional imaging suggests DLL3 as a promising therapeutic target in both GEP NECs and high grade PanNETs.

## INTRODUCTION

Delta-like ligand 3 (DLL3) is an inhibitory Notch pathway ligand and a target of ASCL1^1^, a critical transcription factor driving neuroendocrine cell fate decisions in small cell lung cancer (SCLC)^2^. DLL3 is highly expressed on the cell surface in SCLC and neuroendocrine prostate cancer (NEPC) with minimal expression on normal cells^3,4^.

These observations have led to expanding efforts to target DLL3 in SCLC and NEPC^5^, including DLL3-targeted T cell engagers (TCEs)^6,7,8^, antibody-drug conjugates^9^, and radioligand therapies^10,11^. Most notably, tarlatamab, a DLL3-CD3 bispecific TCE, recently demonstrated benefit over standard of care chemotherapy in patients with previously treated SCLC^12^. In addition, recent work describing functional DLL3 radiotracer uptake in patients with SCLC and NEPC^13^ has prompted exploration of radioligand therapy development in patients with these tumors^10, 11^.

Despite ongoing development of DLL3-targeted therapies in SCLC and NEPC, the potential role for these agents in patients with other extrapulmonary neuroendocrine neoplasms remains underexplored. Gastroenteropancreatic neuroendocrine neoplasms (GEP NENs), which include well differentiated neuroendocrine tumors (GEP NETs) and poorly differentiated neuroendocrine carcinomas (GEP NECs), now represent the second most common gastrointestinal malignancies^14^, and are in need of new therapeutic options. Patients with well differentiated GEP NETs are often treated with somatostatin receptor (SSTR)-targeted therapies, including somatostatin analogs^15, 16^ and SSTR-targeted radioligand therapies (^177^Lu-DOTATATE^17, 18^), as well as targeted therapies^19, 20^, and chemotherapies^21, 22^. However, the optimal treatment for patients with high grade (grade 3 [G3]) GEP NETs, who harbor an intermediate prognosis between that of G1-2 GEP NETs and GEP NECs^23^, remains unknown. These patients are often marked by lower and/or more heterogeneous SSTR PET avidity compared to G1-2 NETs and are often treated with chemotherapies; however, response rates to chemotherapies are generally lower in G3 GEP NETs compared to GEP NECs^24^. In contrast, GEP NECs are almost uniformly treated with platinum-based chemotherapies, although optimal first-line therapy remains uncertain^25^, with minimal role for SSTR targeting^26^. In sum, there is a need for new therapies for both G3 GEP NETs and GEP NECs.

Given the success of SSTR-targeted radioligand therapies in patients with GEP NENs^17, 18^, whether similar opportunities are possible with DLL3- or other cell surface antigen-directed therapies is of key interest, particularly in the G3 GEP NET and GEP NEC populations, in whom benefits of SSTR-targeted radioligand therapies are often limited ^27, 26^. In addition, the degree of DLL3 expression required for actionability of DLL3 targeted therapies, and the degree of DLL3 intertumoral heterogeneity in patients with GEP NENs remain unknown, for which development of a functional, dynamic, patient-level marker of DLL3 expression is required. Here, we describe the landscape of DLL3 IHC expression in patients with GEP NENs and feasibility of [^89^Zr]Zr-DFO-SC16.56 functional DLL3 PET imaging^13^, with the goal of further defining opportunities for DLL3 therapeutic targeting in these populations.

## METHODS

### Patients, tissue samples, and clinicopathologic analyses

All patients with histologically confirmed poorly differentiated GEP NECs and grade 3 well-differentiated GEP NETs treated between January 1, 2018 and June 1, 2025 at Memorial Sloan Kettering Cancer Center (MSK) and with available pathology specimens were identified, with the results of DLL3 IHC performed prospectively as below collected. Patients in whom DLL3 IHC was not performed prospectively were profiled retrospectively through generation of formalin-fixed, paraffin-embedded tissue slides or tissue microarrays (TMAs) created with two cores per sample (core diameter 1.5 mm) from a representative tumor area. DLL3 IHC was also performed on TMAs constructed from surgically resected G1-G2 small bowel, pancreatic, and colorectal NETs treated between 2018-2024, resected G1-G2 pancreatic NETs (PanNETs) treated between 2002-2009, and resected G1 small bowel NETs treated between 1995-2008. For patients with available tissue samples and DLL3 IHC data, sociodemographic characteristics, clinicopathologic information, MSK-IMPACT^28^ next generation sequencing data, results from radiographic assessments, and outcomes to systemic therapies were collected. The data cutoff for retrospective analyses was June 1, 2025. This portion of the study was approved by the Institutional Review Board/Privacy Board at MSK under biospecimen research protocol 16-1228 and was in accordance with the Belmont report for retrospective review of records and waiver of consent.

### DLL3 immunohistochemistry (IHC)

Tumor samples were assessed for DLL3 expression by IHC using a standardized Ventana platform assay (clone SP347, Ventana, Roche, Tucson, AZ)^29^. Semiquantitative assessment of DLL3 expression was based on estimation of the percentage of positive tumor cells (tumor proportion score, range 0-100%) multiplied by staining intensity (weak: 1+, moderate: 2+, strong: 3+) to generate an H-score (range 0-300). Positive DLL3 expression was defined as ≥ 5% of tumor cells with weak (1+) or higher staining. All staining assessments were completed by board-certified pathologists (J.S., O.B., L.H.T. for prospective samples, Z.C.T., U.B. for retrospective samples and TMAs).

### MSK-IMPACT next generation sequencing (NGS)

Briefly, genomic DNA was extracted from patient tumors and matched normal DNA from peripheral blood samples. Barcoded libraries were generated and sequenced, targeting all exons and select introns of a custom panels of 341-505 genes (MSK-IMPACT versions 1-7). Somatic substitutions, small insertions/deletions, gene-level focal copy-number amplifications, homozygous deletions, and fusions in select genes were identified using a clinically validated pipeline^28^.

### Retrospective evaluation of ^68^Ga-DOTATATE PET-CT characteristics

Among patients with PanNETs who underwent ^68^Ga-DOTATATE PET-CT imaging within six months of tumor tissue collection for DLL3 IHC, a board-certified radiologist (L.B.) evaluated ^68^Ga-DOTATATE PET-CT characteristics of tumor lesions at corresponding sites of tumor tissue evaluation. For each patient, maximum standard uptake value (SUV_max_), tumor/liver, and tumor/spleen ratios for the corresponding tumor lesion was identified by evaluating SUV_mean_ for liver and spleen background for each patient when able. Patients who received intercurrent local therapies or had a change in systemic therapy in between tumor tissue collection for DLL3 IHC and ^68^Ga-DOTATATE PET-CT were excluded.

### [^89^Zr]Zr-DFO-SC16.56 DLL3 immunoPET-CT imaging

Between May 28, 2024 and February 10, 2025, selected patients with advanced DLL3 IHC-positive GEP NENs with recent progression of disease (within ≤ 12 weeks) on standard systemic therapy were enrolled onto a phase II study of the humanized anti-DLL3 monoclonal antibody SC16.56, labeled with Zirconium-89 (^89^Zr) for PET imaging^13^. The radioimmunoconjugate [^89^Zr]Zr-DFO-SC16.56 was manufactured at the MSK Radiochemistry and Molecular Imaging Probes Core Facility. SC16.56 anti-DLL3 antibody (AbbVie, Chicago, IL, USA) was conjugated with a bifunctional chelator, p-SCN-Bn-deferoxamine (DFO; Macrocyclics, Plano, TX, USA), and radiolabeled with ^89^Zr-oxalate (3D Imaging, Little Rock, AR, USA). Patients received a single 2.5 mg intravenous injection of [^89^Zr]Zr-DFO-SC16.56, which included 2.5 mg of SC16.56 and 37-74 MBq of ^89^Zr, followed by a PET-CT scan 3-5 days later^13^. Images were analyzed by board-certified nuclear medicine physicians (M.P.D., L.B.) using commercial display software (Hermes Gold4.4-B, AW Centricity Imaging-PACS/AW Suite). Tumoral SUVs were measured from reconstructed PET images, and were expressed in terms of SUV_max_ and SUV_mean_. For quantitative tumor analyses, the lesion with the highest SUV was selected from each patient, with tumors with tracer retention greater than blood pool considered avid. In addition, the number and percentage of DLL3-positive tumor lesions were calculated comparing [^89^Zr]Zr-DFO-SC16.56 DLL3 PET imaging with results of recent standard of care scans including CT or ^68^Ga-DOTATATE PET-CT within ≤ 12 weeks, with inclusion criterion for evaluable tumor lesions including soft tissue lesions > 1.5 cm (defined as limit of DLL3 PET spatial resolution). All patients provided written informed consent under MSK protocol 19-292, which was done in accordance with the Declaration of Helsinki. This study is ongoing and is registered with ClinicalTrials.gov (NCT04199741).

### Statistical analyses

Sociodemographic, clinicopathologic, and treatment characteristics were summarized among DLL3 IHC-positive and negative groups using frequencies and percentages for categorical variables, and median and interquartile ranges for continuous variables. Covariate distributions between DLL3-positive and negative groups were compared using Fisher exact tests for categorical variables and Wilcoxon rank-sum tests for continuous variables.

For genomic enrichment analyses among patients with available DLL3 IHC and MSK-IMPACT NGS data, Fisher exact tests were used to calculate *P* values for differences in frequency of OncoKB^30^ oncogenic alterations between groups; *Q* values were calculated using the false discovery rate/Benjamini Hochberg method and corrected for the number of genes with ≥ 5% oncogenic alterations in each cohort.

For clinical outcome analyses among patients with advanced GEP NECs, advanced G3 PanNETs, and advanced PanNETs (all grades) respectively, overall survival (OS) was calculated from the date of tissue collection, where DLL3 IHC status was ascertained, until the date of last follow up or death. Among patients with advanced GEP NECs, progression-free survival (PFS) to first-line platinum-based chemotherapy was calculated from the date of start of platinum-based chemotherapy until date of first disease progression or death, whichever occurred first. PFS and OS were estimated using Kaplan-Meier methods and compared between groups using log-rank tests. In addition, among advanced G3 PanNETs and advanced PanNETs (all grades), Cox regression models were used to correlate DLL3 IHC status with OS from date of diagnosis of advanced G3 disease or advanced disease of any grade, respectively, incorporating DLL3 IHC status as a time-dependent covariate.

All statistical analyses were performed using R Version 4.3.2 (R Foundation for Statistical Computing, Vienna, Austria). Genomic enrichment analyses were performed using cBioPortal^31^. All *P*-values were two-sided, and *P*-values of < 0.05 were considered statistically significant.

## RESULTS

### Landscape of DLL3 expression in GEP NECs

We first aimed to describe DLL3 expression in poorly differentiated neuroendocrine carcinomas (GEP NECs, **Table 1**). DLL3 expression was detected in 53 of 75 GEP NEC samples overall (71%; median IHC H-score 45, interquartile range [IQR] 0-160, range 0-300); breakdown by GEP NEC subtype is displayed in **Figures 1A-1B** and **Table 1**. Among GEP NECs, DLL3 expression was detected in 21 of 25 samples with small cell (84%), 13 of 21 with large cell histology (62%), and 19 of 29 (66%) samples with histology not otherwise specified; median H-score was significantly higher among GEP NEC tumors with small cell compared to large cell histology (median 120 versus 10, *P* = 0.007, **Figure 1C, Supplementary Table 1**). DLL3 expression was not significantly different between primary versus metastatic samples (**Supplementary Figure 2, Supplementary Table 1**), and median Ki67 proliferative index was not significantly different by DLL3 status (*P* = 0.070, **Supplementary Figure 1A, Supplementary Table 1)**. In addition, among GEP NEC tumors with available MSK-IMPACT NGS (n = 45), there were no significant differences in frequency of individual oncogenic driver alterations among DLL3-positive compared to DLL3-negative tumors overall (**Figure 1D**) or within any GEP NEC subtype (**Supplementary Table 2**).

**Figure 1:**
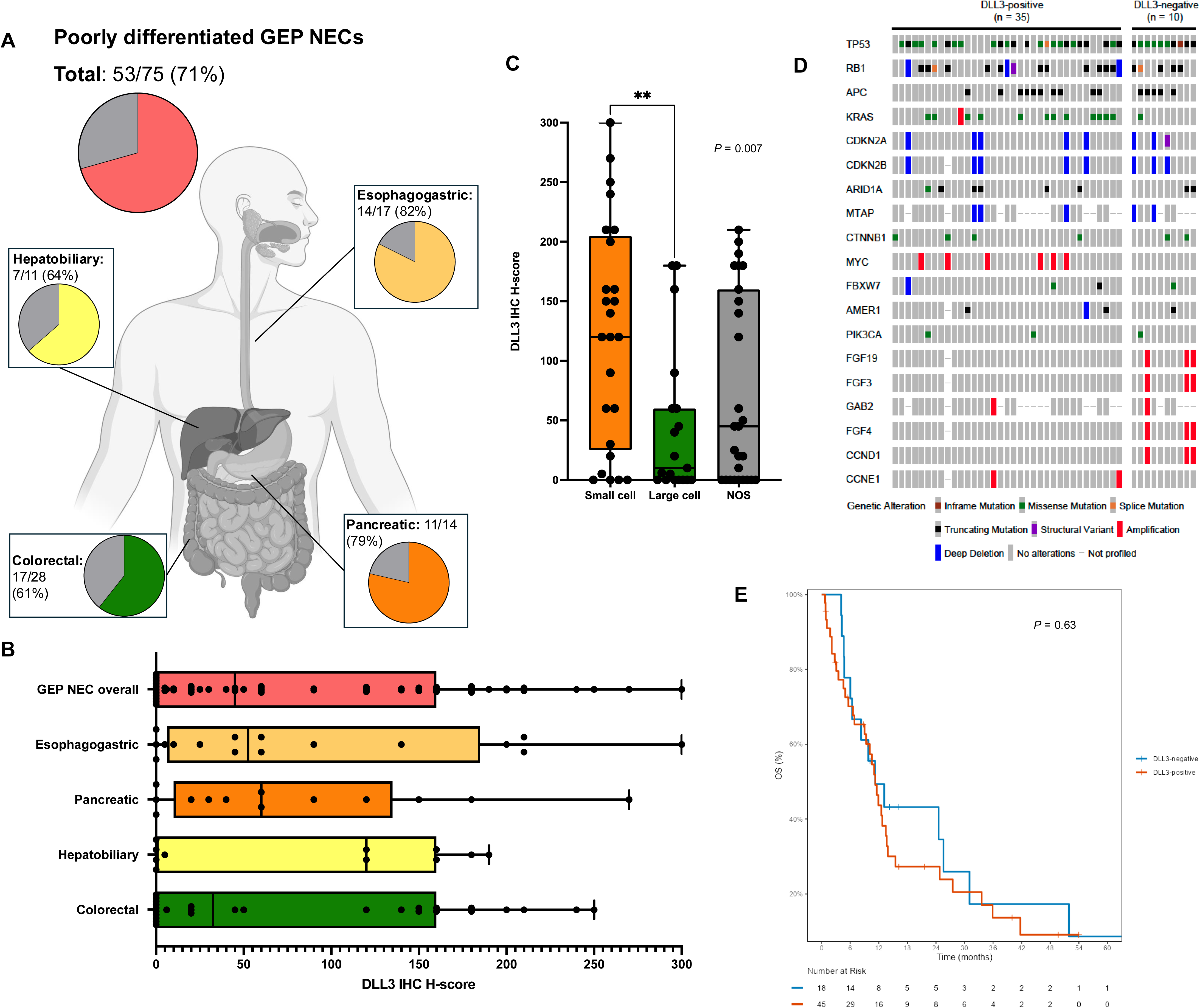
Landscape of DLL3 immunohistochemistry (IHC) expression and clinicopathologic correlates of DLL3 expression in poorly differentiated gastroenteropancreatic neuroendocrine carcinomas (GEP NECs). **(A)** DLL3 IHC positivity rates among GEP NECs overall and by primary site of disease. **(B)** Box and whisker plots describing median, interquartile range, and distribution of individual DLL3 IHC H-scores among tumor samples from poorly differentiated GEP NECs overall and by primary site of origin. **(C)** Box and whisker plots describing median, interquartile range, and distribution of individual DLL3 immunohistochemistry (IHC) H-scores among GEP NEC tumor samples with small cell histology (n = 25), large cell histology (n = 21), or histology not otherwise specified (NOS, n = 29). *P* value refers to comparison between small cell and large cell groups by Wilcoxon rank-sum test. **(D)** OncoPrint of genomic alterations by DLL3 status among GEP NEC tumors with available DLL3 IHC and MSK-IMPACT next generation sequencing (NGS). Only genes with at least 5% prevalence of oncogenic or likely oncogenic alterations in the cohort are displayed. **(E)** Overall survival (OS) from diagnosis of advanced disease by DLL3 IHC status among patients with advanced GEP NECs. *P* value corresponds to log-rank test.

**Table 1:** DLL3 immunohistochemistry (IHC) expression in gastroenteropancreatic neuroendocrine neoplasms. Displayed are DLL3 IHC positivity rates among poorly differentiated GEP neuroendocrine carcinomas (GEP NECs) overall and by primary site of disease, and well differentiated pancreatic (PanNETs), small bowel (SB NETs), and colorectal NETs by grade (G1, G2, and G3). Other GEP NEC included small bowel NEC (n = 3) and NEC of unknown primary (n = 2). IHC positivity was defined as ≥ 5% of tumor cells with ≥ weak (1+) IHC staining. IQR = interquartile range.

|  | **DLL3-positive (%)** | **DLL3 IHC H-score**<br>**(median, IQR)** |
| --- | --- | --- |
| **GEP NEC overall** | 53/75 (71%) | 45 (0, 160) |
| Esophagogastric | 14/17 (82%) | 53 (6, 185) |
| Pancreatic | 11/14 (79%) | 60 (10, 135) |
| Hepatobiliary | 7/11 (64%) | 120 (0, 160) |
| Colorectal | 17/28 (61%) | 33 (0, 160) |
| Other | 4/5 (80%) | 20 (5, 180) |
| **Pancreatic NET (PanNET)** |  |  |
| G1 PanNET | 3/46 (7%) | 0 (0, 1) |
| G2 PanNET | 1/23 (4%) | 0 (0, 1) |
| G3 PanNET | 22/51 (43%) | 0 (0, 30) |
| **Small bowel NET (SB NET)** |  |  |
| G1 SB NET | 0/137 (0%) | 0 (0, 0) |
| G2 SB NET | 1/21 (5%) | 0 (0, 0) |
| G3 SB NET | 1/5 (20%) | 0 (0, 10) |
| **Colorectal NET** |  |  |
| G1 colorectal NET | 0/5 (0%) | 0 (0, 0) |
| G2 colorectal NET | 0/2 (0%) | 0 (0, 0) |
| G3 colorectal NET | 2/5 (40%) | 0 (0, 39) |

We next sought to determine the prognostic significance of DLL3 expression in patients with advanced GEP NECs (n = 64, **Supplementary Table 3**). Among patients treated with first-line platinum-based chemotherapy (n = 37), PFS was not significantly different between DLL3-positive tumors compared to DLL3-negative tumors (median 2.9 months [mo], 95% confidence interval [CI] 1.9-9.6 mo versus median 4.3 mo, 95% CI 2.3 mo-not reached [NR], *P* = 0.57, **Supplementary Figure 3**). Similarly, OS was not significantly different between DLL3-positive compared to DLL3-negative tumors (median OS 11 mo, 95% CI 9-14 mo versus 11 mo, 95% CI 6.4 mo-NR, *P* = 0.63, **Figure 1E**).

### Landscape of DLL3 expression in well differentiated GEP NETs

We next aimed to describe DLL3 expression in well differentiated GEP NETs by grade and primary site of origin, including pancreatic (PanNET), small bowel, and colorectal NETs. Among 120 PanNETs tested, DLL3 expression was almost exclusively restricted to G3 tumors, with 22 of 51 (43%) G3 tumors positive for DLL3 expression (median IHC H-score 0, IQR 0-30, range 0-270; **Figures 1A-1B**, **Table 1**). DLL3 expression was detected in 1 of 5 (20%) G3 small bowel NETs (**Figure 1A**) and 2 of 5 (40%) G3 colorectal NETs (**Table 1**); in total, 25 of 61 (41%) G3 GEP NETs tested positive for DLL3 expression.

Given that G3 pancreatic NETs are more commonly observed in clinical practice^23^, we next aimed to specifically define the landscape of DLL3 expression in PanNETs. Median Ki67 was significantly higher in DLL3-positive compared to DLL3-negative tumors among PanNETs overall (45% versus 7%, *P* < 0.001, **Supplementary Figure 1B**) and within G3 PanNETs specifically (49% versus 30%, *P* = 0.005, **Figure 2C**). Among G3 PanNETs, there was no association between DLL3 positivity and frequency of individual oncogenic alterations (**Figure 2D, Supplementary Table 4**). Next, given the clear association of DLL3 positivity with high grade disease (**Figures 1A-1C, Supplementary Figure 1B**), we aimed to explore whether DLL3 tumor IHC positivity was inversely correlated with SSTR PET avidity. Interestingly, among 24 patients with PanNETs who underwent ^68^Ga-DOTATATE PET-CT imaging within six months of tumor tissue collection for DLL3 IHC, there was no association between tumor DLL3 IHC positivity and corresponding tumor lesion SUV_max_, tumor/liver ratio, or tumor/spleen ratio on ^68^Ga-DOTATATE PET-CT (**Supplementary Figures 4A-4C**), suggesting that DLL3 expression in high grade PanNETs may not be mutually exclusive with SSTR PET avidity.

**Figure 2:**
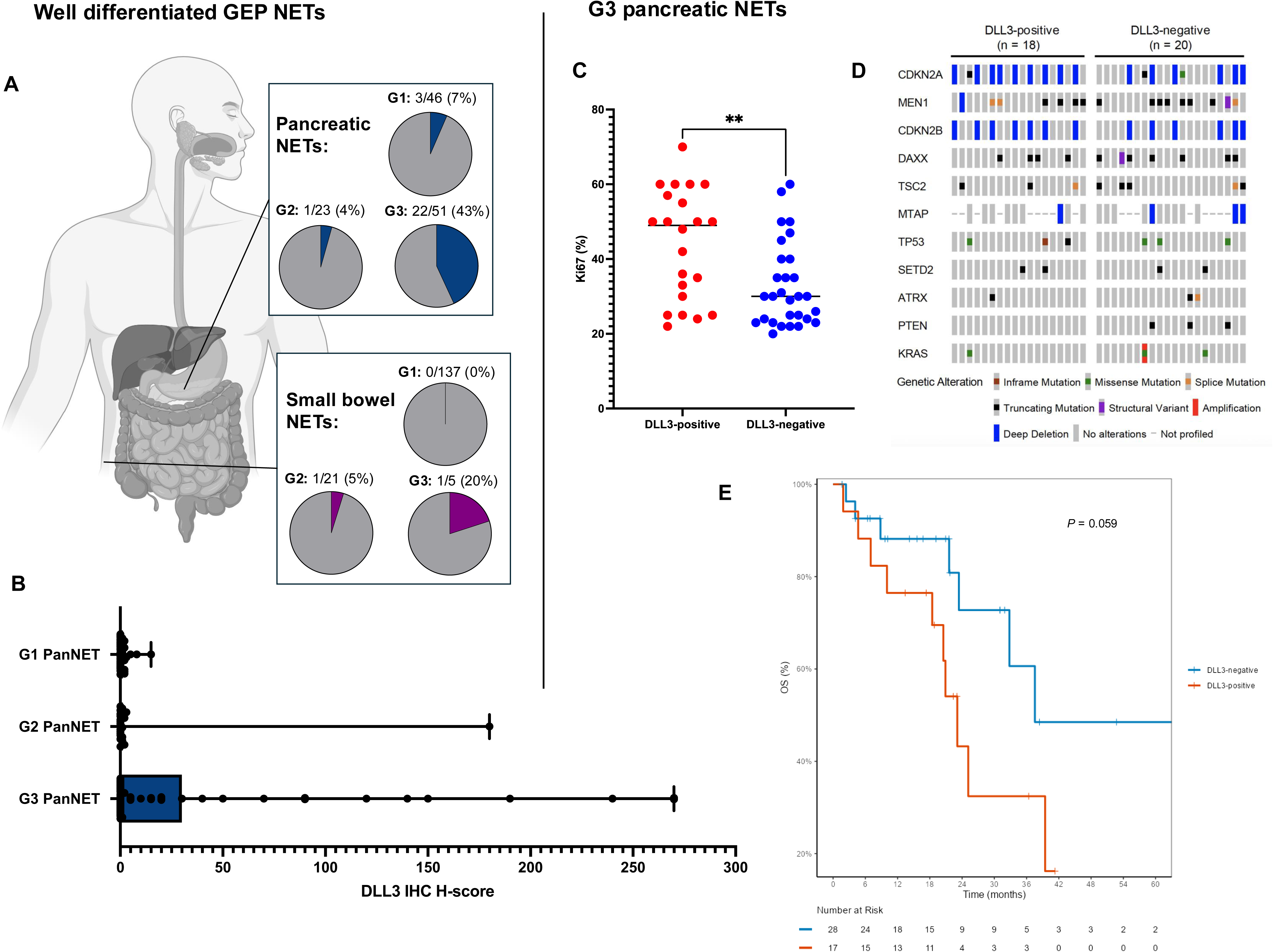
Landscape of DLL3 immunohistochemistry (IHC) expression in well differentiated gastroenteropancreatic neuroendocrine tumors (GEP NETs) and clinicopathologic correlates of DLL3 expression in grade 3 (G3) well differentiated pancreatic NETs. **(A)** DLL3 IHC positivity rates among well differentiated pancreatic neuroendocrine tumors (PanNETs) and small bowel NETs by grade (G1: grade 1, G2: grade 2, G3: grade 3). **(B)** Box and whisker plots describing median, interquartile range, and distribution of individual DLL3 IHC H-scores among tumor samples from PanNETs by grade. **(C)** Distribution of Ki67 percentages (%) among G3 PanNETs by DLL3 (IHC) positive (n = 22) and negative (n = 29) status. *P* value refers to comparison between groups by Wilcoxon rank-sum test. **(D)** OncoPrint of genomic alterations by DLL3 status among G3 PanNET tumors with available DLL3 IHC and MSK-IMPACT NGS. Only genes with at least 5% prevalence of oncogenic or likely oncogenic alterations in the cohort are displayed. **(E)** Overall survival (OS) from time of tumor evaluation by DLL3 IHC in patients with advanced G3 PanNETs. *P* value corresponds to log-rank test.

Finally, given association of DLL3 positivity with high grade disease, we aimed to understand the prognostic significance of DLL3 tumor positivity among patients diagnosed with advanced PanNETs (n = 66, **Supplementary Table 5**). We observed shorter OS in patients diagnosed with DLL3-positive G3 PanNETs (*P* = 0.059, **Figure 2E**) as well in DLL3-positive advanced PanNETs of all grades (*P* < 0.001, **Supplementary Figure 5A**), with systemic therapy regimens generally well balanced between the DLL3-positive and DLL3-negative groups (**Supplementary Table 5**). Understanding that timing of biopsies may be variable in patients’ disease courses, we also observed trend toward shorter OS among DLL3-positive patients when restricting to patients in whom DLL3 status was evaluated at initial diagnosis of G3 advanced disease (n = 35, *P* = 0.094, **Supplementary Figure 5B**). In addition, using a Cox regression model incorporating DLL3 IHC status as a time-dependent covariate, we observed significantly increased risk for mortality from time of diagnosis of G3 advanced disease among DLL3-positive patients (OS hazard ratio [HR] 3.27, 95% CI 1.09-9.78, *P* = 0.034). Similar results were observed when considering advanced PanNETs of all grades, with Cox regression analysis demonstrating an OS HR from diagnosis of advanced disease of 4.53 (95% CI 1.86-11.0, *P* < 0.001).

### DLL3 immunoPET-CT imaging reveals DLL3 as an actionable target in GEP NENs

To evaluate whether DLL3 tumor IHC positivity in GEP NENs is associated with functional expression and potential for therapeutic targeting, we completed [^89^Zr]Zr-DFO-SC16.56 immuno-PET-CT imaging^13^ on six patients with advanced DLL3 IHC-positive GEP NENs (**Table 2, Supplementary Table 6**) as part of a phase II PET imaging trial. In total, tumor-specific [^89^Zr]Zr-DFO-SC16.56 uptake was observed in five of six patients, including both patients with GEP NECs and three of four patients with G3 PanNETs, with minimal background uptake (**Table 2**, **Figures 3-4, Supplementary Figure 6**). No adverse events were observed (**Supplementary Table 6**).

**Figure 3:**
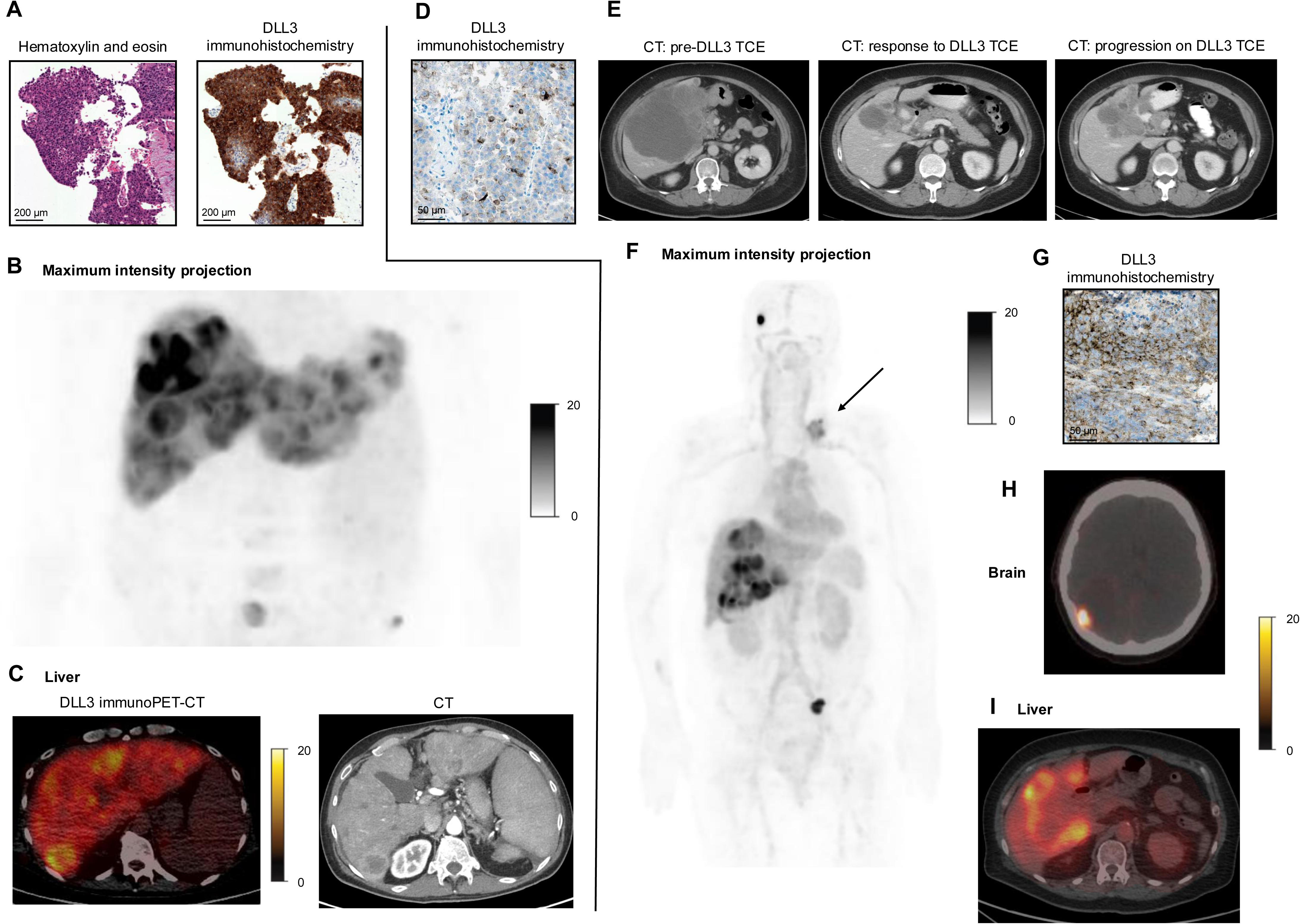
[^89^Zr]Zr-DFO-SC16.56 DLL3 immunoPET-CT imaging in patients with advanced poorly differentiated gastroenteropancreatic neuroendocrine carcinomas (GEP NECs). **(A-C):** A patient with poorly differentiated pancreatic neuroendocrine carcinoma (NEC) metastatic to liver with prior progression of disease on carboplatin and etoposide and CAPOX underwent [^89^Zr]Zr-DFO-SC16.56 DLL3 immunoPET-CT imaging (patient 1). **(A)** Hematoxylin and eosin (H&E) and DLL3 immunohistochemistry (IHC) images from a liver metastasis biopsy prior to start of systemic therapy are shown, revealing DLL3 IHC H-score of 270. **(B)** Maximum intensity projection (MIP) and **(C)** axial images from DLL3 immunoPET-CT are shown, with corresponding CT completed two days after DLL3 immunoPET-CT shown. **(D-I):** A patient with gallbladder NEC with multiple prior lines of systemic therapy underwent [^89^Zr]Zr-DFO-SC16.56 DLL3 immunoPET-CT imaging after prior progression of disease on DLL3 T cell engager therapy (patient 2). **(D)** DLL3 IHC from a liver metastasis at biopsy confirmation of NEC histology prior to DLL3 TCE therapy revealed IHC H-score 160. **(E)** Serial CT scans on DLL3 TCE therapy revealing partial response followed by progression of disease. At progression of disease on fourth line carboplatin/etoposide, [^89^Zr]Zr-DFO-SC16.56 DLL3 immunoPET-CT was completed, with **(F)** MIP and **(H-I)** axial images highlighting brain and liver metastases shown. **(G)** Biopsy of DLL3 PET-avid left supraclavicular nodal metastasis (corresponding to arrow in MIP image) revealed retained DLL3 IHC expression (H-score 120). For MIP and axial images respectively, standard uptake value (SUV) scales are displayed. For MIP images, pixels in PET imaging with SUV 0 appear white and pixels with SUV ≥ upper thresholds as shown appear as black. For axial images, pixels with SUV 0 and SUV ≥ upper thresholds as shown appear black and bright yellow respectively.

**Figure 4:**
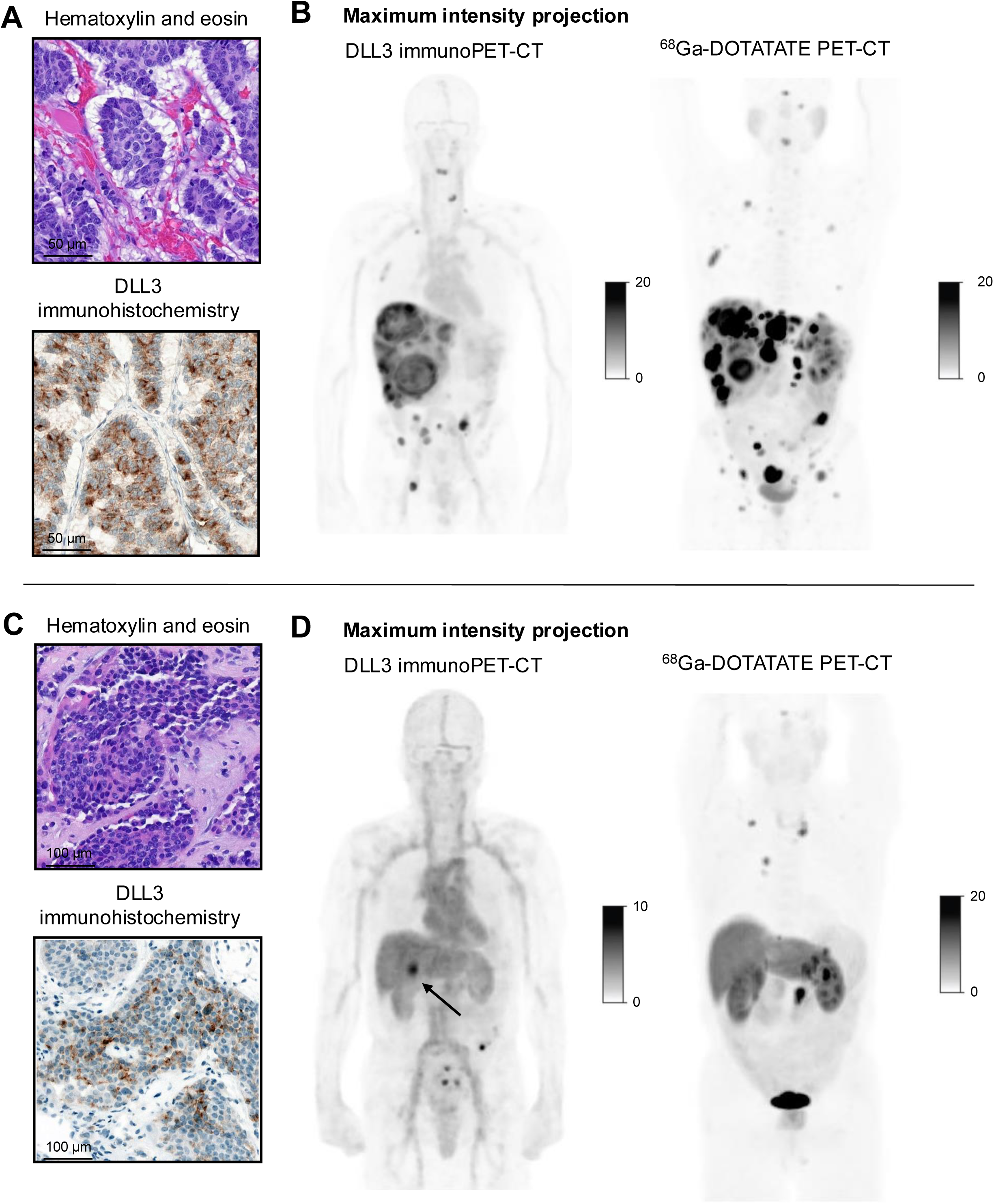
[^89^Zr]Zr-DFO-SC16.56 DLL3 immunoPET-CT imaging in patients with advanced grade 3 well differentiated pancreatic neuroendocrine tumors (G3 PanNETs). **(A-B)** A patient with G3 PanNET metastatic to liver underwent [^89^Zr]Zr-DFO-SC16.56 DLL3 immunoPET-CT imaging at progression of disease on third-line ^177^Lu-DOTATATE (patient 5). **(A)** H&E and DLL3 immunohistochemistry (IHC) images from the patient’s pancreatic primary tumor prior to start of systemic therapy are shown. **(B)** Maximum intensity projection (MIP) images from DLL3 immunoPET-CT and ^68^Ga-DOTATATE PET-CT imaging completed 12 weeks prior to DLL3 immunoPET-CT are shown, revealing 100% of tumor lesions positive for both DLL3 and somatostatin receptor avidity. **(C-D)** A patient with G3 PanNET metastatic to liver, lung, and mediastinum underwent DLL3 immunoPET-CT after progression of disease in the liver on second-line capecitabine and temozolomide. **(C)** H&E and DLL3 IHC images from progressive liver metastases are shown, revealing DLL3 IHC 120. (**D)** MIP images from DLL3 immunoPET-CT and ^68^Ga-DOTATATE PET-CT imaging completed 5 weeks prior to DLL3 immunoPET-CT are shown, revealing DLL3 uptake in the progressive liver metastasis (SUV_max_ 14.4); the remainer of the disease was otherwise DLL3 PET-negative and SSTR PET-positive. Arrow notes progressive liver metastatic lesion which was biopsied for DLL3 IHC evaluation. For all images, standard uptake value (SUV) scales are displayed, with pixels in PET imaging with SUV 0 appearing white and pixels with SUV ≥ upper thresholds as shown appearing black.

**Table 2:** Patient characteristics and results of [^89^Zr]Zr-DFO-SC16.56 DLL3 immunoPET-CT imaging. NEC = neuroendocrine carcinoma. G3 PanNET = grade 3 well differentiated pancreatic neuroendocrine tumor. IHC = immunohistochemistry. SUV_max_ = maximum standard uptake value.

| Patient # | Tumor type | DLL3 IHC H-score (biopsy location) | Number of prior lines of systemic therapy for advanced disease | Most recent systemic therapy prior to DLL3 immuno-PET CT | DLL3 immuno PET-CT avid | SUV<sub>max</sub> of highest DLL3-avid lesion (location) | Blood pool uptake, SUV<sub>mean</sub> | Tumor/liver ratio, SUV<sub>max</sub>/SUV<sub>mean</sub> | # (%) of tumor lesions DLL3 immunoPET-CT positive | Among DLL3 immunoPET-CT positive tumor lesions, % <sup>68</sup>Ga-DOTATATE positive (PanNET only) |
| --- | --- | --- | --- | --- | --- | --- | --- | --- | --- | --- |
| 1 | Pancreatic NEC | 270 (liver) | 2 | CAPOX | Yes | 36.7 (liver) | 1.6 | 5.7 | 20/21 (95%) | N/A |
| 2 | Gallbladder NEC | 120 (supra-clavicular lymph node) | 4 | Carboplatin + etoposide | Yes | 32.4 (liver) | 7.9 | 4.8 | 16/16 (100%) | N/A |
| 3 | G3 PanNET | 90 (liver) | 4 | Capecitabine + temozolomide | No | 7.4 (abdominal lymph node) | 7.9 | 1.1 | 0/22 (0%) | N/A |
| 4 | G3 PanNET | 120 (liver) | 2 | Capecitabine + temozolomide | Yes | 14.4 (liver) | 4.8 | 3.1 | 1/10 (10%) | 0% |
| 5 | G3 PanNET | 90 (pancreatic primary) | 3 | <sup>177</sup>Lu-DOTATATE | Yes | 27.5 (liver) | 3.9 | 5.2 | 38/38 (100%) | 100% |
| 6 | PanNET | 30 (liver) | 3 | CAPOX | Yes | 15.0 (liver) | 5.5 | 2.7 | 4/8 (50%) | 100% |

### DLL3 immunoPET-CT imaging in patients with GEP NECs

Two patients with GEP NECs were imaged. Patient 1 (**Figures 3A-3C**) was a patient with advanced pancreatic small cell NEC who experienced recent progression on second-line chemotherapy with DLL3 IHC from a progressing tumor lesion demonstrating H-score of 270 (**Figure 3A**). High level tumor-specific [^89^Zr]Zr-DFO-SC16.56 uptake was demonstrated, with DLL3 PET SUV_max_ of 36.7 and 95% of tumor lesions demonstrating DLL3 PET avidity (**Figures 3B-3C**). Patient 2 (**Figures 3C-3I**) was initially diagnosed with a primary gallbladder carcinoma and experienced rapid progression of disease on two lines of platinum-based chemotherapy. At progression on second-line therapy, cell free DNA profiling through an institutional platform^32^ revealed pathogenic *TP53*, *RB1* mutations and *MYC* amplification, with repeat liver lesion biopsy revealing high grade neuroendocrine carcinoma, DLL3 IHC H-score 160 (**Figure 3D**).

As a result, the patient was treated with a DLL3 bispecific T cell engager (TCE) with clinical and radiographic response (**Figure 3E**). The patient ultimately developed progressive disease on DLL3 TCE therapy, prompting switch in therapy. At further progression, the patient underwent [^89^Zr]Zr-DFO-SC16.56 DLL3 PET imaging which revealed avidity in all active tumor lesions, with highest uptake observed in a progressing liver metastasis (SUV_max_ 32.4, **Figure 3F**, **Figures 3H-3I**). A progressive left supraclavicular lymph node demonstrated SUV_max_ 10.9, prompting biopsy revealing DLL3 IHC H-score 120 (**Figure 3G**). In addition, a DLL3-avid asymptomatic brain metastasis was identified (SUV_max_ 29.1, **Figure 3H**). Given confirmation of uniformly retained high level DLL3 PET avidity after progression on prior DLL3 TCE therapy, the patient was considered for treatment with a DLL3-directed antibody-drug conjugate.

### DLL3 immunoPET-CT imaging in patients with high grade pancreatic NETs

DLL3 immunoPET-CT imaging was also completed in patients with well differentiated PanNETs, with degree of tracer avidity and interlesional heterogeneity more variable. Four patients with DLL3 IHC-positive advanced G3 PanNETs were imaged (IHC H-score range: 30-120), with three of four demonstrating [^89^Zr]Zr-DFO-SC16.56 uptake, ranging from SUV_max_ 14.4 to 27.5, with 10% to 100% of tumor lesions demonstrating DLL3 PET avidity (**Table 2**). These included a patient with G3 PanNET with recent progression of disease on ^177^Lu-DOTATATE and prior DLL3 IHC H-score 90, in whom DLL3 immunoPET-CT revealed 100% of active tumor lesions positive for DLL3 tracer uptake with SUV_max_ 27.5 in a progressing liver lesion (patient 5; **Table 2**, **Figure 4A->4B**). In two of three patients (patients 5 and 6, **Figures 4A-4B, Supplementary Figure 6B**), DLL3 immunoPET-CT imaging revealed that all DLL3 PET-positive tumor lesions also demonstrated SSTR avidity at the individual tumor level on corresponding ^68^Ga-DOTATATE PET-CT imaging. In contrast, we observed one patient with advanced G3 PanNET and recent progression of disease in the liver on capecitabine and temozolomide (patient 4, **Figure 4C-4D**), with a prior liver metastasis biopsy revealing DLL3 IHC H-score 120 (**Figure 4C**). In this patient, the progressive liver lesion was positive on [^89^Zr]Zr-DFO-SC16.56 PET imaging (SUV_max_ 14.4) but was not visualized on concurrent SSTR imaging, whereas the remainder of the disease was DLL3 PET-negative and SSTR PET-positive (**Figure 4D**). Overall, these suggested DLL3 avidity can be observed both alongside and independently of SSTR avidity in patients with PanNETs.

## DISCUSSION

Here, using immunohistochemistry and functional PET imaging, we describe the clinical landscape of DLL3 expression in GEP NENs. We demonstrate the potential for DLL3-directed therapy development in poorly differentiated GEP NECs and subsets of well differentiated GEP NETs, particularly high grade pancreatic NETs, and describe the potential utility of [^89^Zr]Zr-DFO-SC16.56 DLL3 immunoPET-CT imaging to guide therapy selection in these patients.

Our demonstration of DLL3 IHC expression in a majority of GEP NECs adds to the growing body of literature describing the prevalence of DLL3 expression in these tumors. Similar recent efforts have estimated DLL3 expression ranging from 44% to 77% in GEP NECs^33, 34, 35^; notably, thresholds for DLL3 IHC positivity have varied across studies. In concert with a recent studies^34, 35^, we observed association of DLL3 expression with small cell histology and identified no prognostic significance for DLL3 expression. Given high prevalence of DLL3 expression, our data provide support for ongoing DLL3 therapeutic trials in GEP NECs^36, 37^ and provide rationale for further drug development in patients with these tumors.

In contrast to GEP NECs, prior studies have reported low DLL3 expression in well differentiated GEP NETs^33, 38^, suggesting minimal role for DLL3 targeting. However, prior efforts have been limited by lack of stratification by tumor grade, primary site, and/or limited inclusion of G3 tumors. Here, we demonstrate DLL3 expression in 43% of G3 pancreatic NETs, and show an association between DLL3 expression and poor outcomes. In concert, two recent studies noted DLL3 expression in 16%^35^ and 53%^34^ of G3 GEP NETs respectively, with minimal expression in G1 and G2 tumors^34, 35^. Together, our data identify DLL3-expressing G3 PanNETs as a robust subset of well differentiated GEP NETs marked by poor outcomes in which DLL3 therapy development should be highly considered.

Our demonstration of robust DLL3 uptake by [^89^Zr]Zr-DFO-SC16.56 immunoPET-CT imaging further supports the role for DLL3 targeted therapy development in GEP NENs. Through therapeutic efforts across NENs, however, the degree of DLL3 IHC expression required for actionability has remained uncertain. As above, varied cutoffs for positive^33, 34, 35, 39^ and high level^34^ expression have been proposed across NENs; in addition, the clinical relevance of membranous versus cytoplasmic DLL3 localization^39^ remains unknown. Furthermore, responses have been observed to tarlatamab in patients with SCLC and no DLL3 IHC expression^6^. Ultimately, DLL3 PET imaging may have better potential to guide treatment selection and stratify patients using real-time functional assessment of DLL3 status across all sites of disease, rather than relying on single site biopsies, which may not reflect DLL3 expression heterogeneity and interval changes over time. In addition, although limited by sample size, it is noteworthy that the three patients with the highest tumor uptake of DLL3 PET tracer in our study had variable DLL3 IHC H-scores of 270, 120, and 90 respectively (**Table 2**), bringing into question the role of IHC from a single tumor tissue sample as a marker for treatment selection.

Our results also suggest that DLL3 immunoPET-CT imaging may be a useful marker for both treatment selection and prognostication in pancreatic NETs. The three cases of DLL3 IHC-positive G3 PanNETs with [^89^Zr]Zr-DFO-SC16.56 PET tracer uptake in our study included a patient with high level DLL3 tracer uptake across all tumor lesions (patient 5, **Figures 4A-4B**), a patient with moderate level uptake in a subset of lesions (patient 6, **Supplementary Figure 6B**), both with concurrent SSTR avidity at all corresponding tumor lesions, and a patient with a DLL3-postive, SSTR-negative escape lesion in the liver with the remainder of the disease otherwise DLL3-negative and SSTR-positive (patient 4, **Figures 4C-4D**). As radioimmunoconjugates targeting DLL3 can be readily converted into therapeutic agents by substituting the imaging isotope with therapeutic radionuclides including Lutetium-177^10, 11^ or Actinium-225^10^, our findings that tumor DLL3 expression and SSTR PET avidity may not be mutually exclusive invoke the intriguing possibility of either sequential or combinatorial targeting with DLL3 and SSTR radioligand therapies in PanNETs guided by functional imaging. As DLL3-expressing PanNET tumors appear to be associated with high grade disease and poor outcomes, targeting DLL3-positive tumor lesions in PanNETs, even in cases of non-uniform expression across tumor sites, may be a promising therapeutic strategy to address higher grade, aggressive tumor subclones that may be associated with poor outcomes.

Finally, given multiple classes of DLL3-targeted therapies in development, including TCEs^6,7,8^, antibody-drug conjugates^9^, and radioligands^10,11^, understanding mechanisms of response and acquired resistance to each class of therapies is critical. Correspondingly, whether patients with prior exposure to a DLL3-targeted therapy may benefit from rechallenge with a DLL3-targeted therapy of a different class after progression of disease remains unclear, as in theory, resistance could proceed through selection for DLL3-negative tumor subclones, or other tumor-intrinsic or extrinsic mechanisms. In our patient with acquired resistance to a prior DLL3 TCE, high level functional DLL3 PET avidity was retained at all sites of disease at progression (**Figures 3F-3I**), suggesting a role for rechallenge with a different class of DLL3-directed therapy. Further work by each class of agents is needed to explore these concepts, with DLL3 immunoPET-CT imaging potentially of value for patient selection in these scenarios.

Our study had a few limitations. This was a single center study with a limited number of [^89^Zr]Zr-DFO-SC16.56 PET scans performed in patients with GEP NENs to date. For biomarker analyses, we included DLL3 IHC data from whole slides and tissue microarrays (TMAs) to increase power. Although a potential limitation, we note that excellent DLL3 IHC concordance between slides and TMA blocks has been demonstrated in prior similar studies^34^. In addition, our ability to retrospectively evaluate DLL3 IHC as a prognostic marker in PanNETs was limited by non-uniform treatment strategies across patients, including different sequencing of systemic therapies and non-uniform administration of local therapies. Recognizing these limitations, however, the association of DLL3 positivity and shorter OS was retained through multiple sensitivity analyses.

In summary, we describe frequent DLL3 IHC expression in high grade pancreatic NETs and poorly differentiated GEP NECs, nominating these tumor types as key populations for development of DLL3-targeted therapies. Our results demonstrate the ability of [^89^Zr]Zr-DFO-SC16.56 PET imaging to successfully delineate DLL3-expressing tumors in patients with GEP NENs, underscoring its potential as a targeted diagnostic tool. As further therapeutic efforts progress, functional DLL3 PET imaging may emerge as comprehensive biomarker for patient selection for novel DLL3-targeted therapies, including radioligand therapies, which have the potential to improve outcomes in patients with high grade GEP NENs.

## Data Availability

All data produced in the present study are available upon reasonable request to the authors.
All data produced in the present work are contained in the manuscript.

## Acknowledgements/Funding

This work was supported in part by the National Institutes of Health (NIH; R01CA213448 [J.T.P.], R35 CA263816 [C.M.R.] and R35 CA232130 [J.S.L.]); a NANETS Clinical Investigator Scholarship (R.T.), and a Prostate Cancer Foundation TACTICAL Award (J.S.L.). We thank AbbVie (Chicago, IL, USA) for generously providing the antibody, SC16.56, for the DLL3 immunoPET-CT portion of the study. Finally, the authors gratefully acknowledge the Memorial Sloan Kettering Molecular Cytology Core and the Tri-Institutional Laboratory of Comparative Pathology, all supported by NIH P30 CA08748.

## FIGURE AND TABLE CAPTION

**Supplementary Figure 1: Correlation of DLL3 immunohistochemistry expression with Ki67 by tumor type.** Distribution of Ki67 percentages (%) among **(A)** poorly differentiated gastroenteropancreatic neuroendocrine carcinomas (GEP NECs) and **(B)** well differentiated pancreatic NETs (PanNETs), including grade 1 to grade 3 disease, by DLL3 expression status. *P* value refers to comparison between groups by Wilcoxon rank-sum test; ns = non-significant.

**Supplementary Figure 2: Correlation of DLL3 immunohistochemistry expression by tumor site among gastroenteropancreatic neuroendocrine carcinomas (GEP NECs).** Comparison of DLL3 IHC H-score distributions among samples from primary tumors (n = 34) versus metastatic disease sites (n = 41). *P* value refers to comparison between groups by Wilcoxon rank-sum test; ns = non-significant.

**Supplementary Figure 3: Progression-free survival (PFS) by DLL3 IHC status among patients with advanced gastroenteropancreatic neuroendocrine carcinomas (GEP NECs) treated with first-line (1L) platinum-based chemotherapy.** Patients who had disease progression (n = 15) prior to DLL3 IHC or DLL3 IHC completed after last available cross-sectional imaging (n = 3) were excluded. *P* value corresponds to log-rank test.

**Supplementary Figure 4: Correlation of DLL3 immunohistochemistry (IHC) expression with ^68^Ga-DOTATATE PET-CT imaging features among patients with pancreatic NETs.** DLL3 IHC was performed on 24 tumor samples in patients with pancreatic NETs (G1: 4, G2: 3, G3: 17; DLL3-positive: 10, DLL3-negative: 14) in whom ^68^Ga-DOTATATE PET-CT imaging was completed within six months of tumor tissue evaluation by DLL3 IHC with no intercurrent local therapies or interim changes in systemic therapy. **(A)** Maximum standard uptake values (SUV_max_), **(B)** Tumor/liver (SUV_max_/SUV_mean_), and **(C)** tumor/spleen (SUV_max_/SUV_mean_) ratios from corresponding index tumor lesions on ^68^Ga-DOTATATE PET-CT imaging are displayed for the DLL3 IHC-positive and IHC-negative groups. *P* values refer to comparisons between groups by Wilcoxon rank-sum test; ns = non-significant.

**Supplementary Figure 5: Prognostic significance of DLL3 expression in patients with advanced well differentiated pancreatic neuroendocrine tumors (PanNETs). (A)** Overall survival (OS) from time of tumor evaluation by DLL3 immunohistochemistry (IHC) in patients with advanced grade 3 (G3) PanNETs, including only patients in whom DLL3 IHC status was available at initial diagnosis of G3 advanced disease. *P* value corresponds to log-rank test. **(B)** OS from time of tumor evaluation by DLL3 IHC in patients with advanced PanNETs (all grades). *P* value corresponds to log-rank test.

**Supplementary Figure 6: Maximum intensity projection (MIP) images from [^89^Zr]Zr-DFO-SC16.56 DLL3 immunoPET-CT imaging in additional patients with advanced well differentiated pancreatic neuroendocrine tumors (PanNETs).** Shown are two additional patients with grade 3 (G3) PanNETs who underwent DLL3 immunoPET-CT imaging. **(A)** Patient 3 underwent DLL3 immunoPET-CT after progression of disease on fourth-line capecitabine and temozolomide. MIP images from ^68^Ga-DOTATATE PET-CT imaging completed six weeks prior to DLL3 immunoPET-CT are shown. **(B)** A patient G3 PanNET metastatic to liver underwent DLL3 immunoPET-CT imaging at progression of disease on third-line CAPOX (patient 6). MIP and axial images from DLL3 immunoPET-CT and ^68^Ga-DOTATATE PET-CT imaging completed seven weeks prior to DLL3 immunoPET-CT are shown. For MIP and axial images respectively, standardized uptake value (SUV) scales are displayed, with pixels in PET imaging with SUV 0 appearing white and black respectively, and pixels with SUV ≥ upper thresholds as shown appearing black and bright yellow respectively.

**Supplementary Table 1:** Correlation of DLL3 immunohistochemistry (IHC) expression with pathologic features among poorly differentiated gastroenteropancreatic neuroendocrine carcinomas (GEP NECs). NOS = not otherwise specified. IQR = interquartile range.

| **Histology** | **DLL3-positive (%)** | **DLL3-negative (%)** | **DLL3 IHC H-score (median, IQR)** |
| --- | --- | --- | --- |
| Small cell (n = 25) | 21 (84%) | 4 (16%) | 120 (25, 205) |
| Large cell (n = 21) | 13 (62%) | 8 (38%) | 10 (0, 60) |
| NOS (n = 29) | 19 (67%) | 9 (66%) | 45 (0, 160) |
| **Site of tissue collection** |  |  |  |
| Primary (n = 34) | 23 (68%) | 11 (32%) | 60 (0, 155) |
| Metastasis (n = 41) | 30 (73%) | 11 (27%) | 45 (10, 160) |
| **Ki67% (Median, IQR)** | 90 (70, 95) | 73 (64, 94) | N/A |

**Supplementary Table 2:**
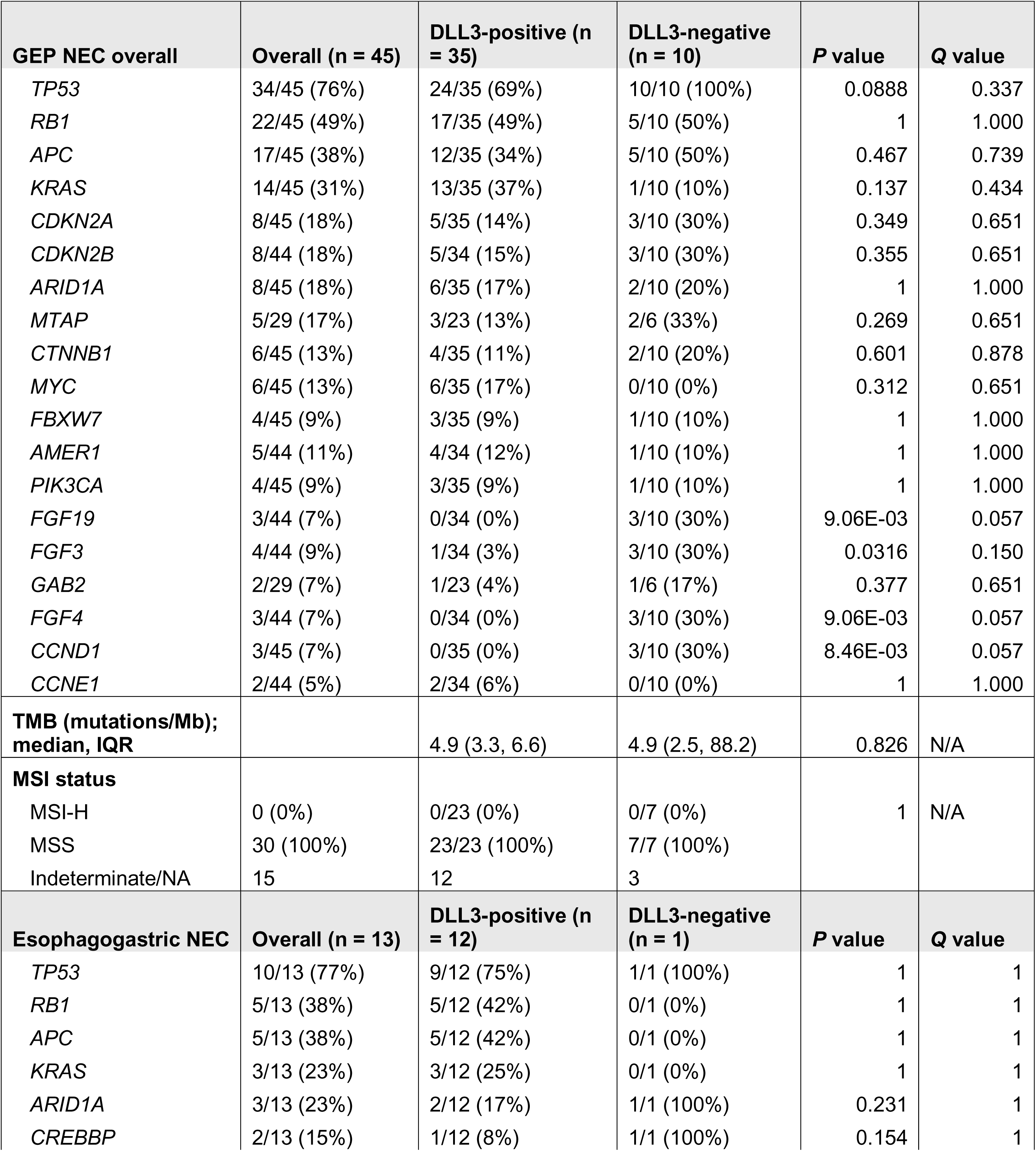
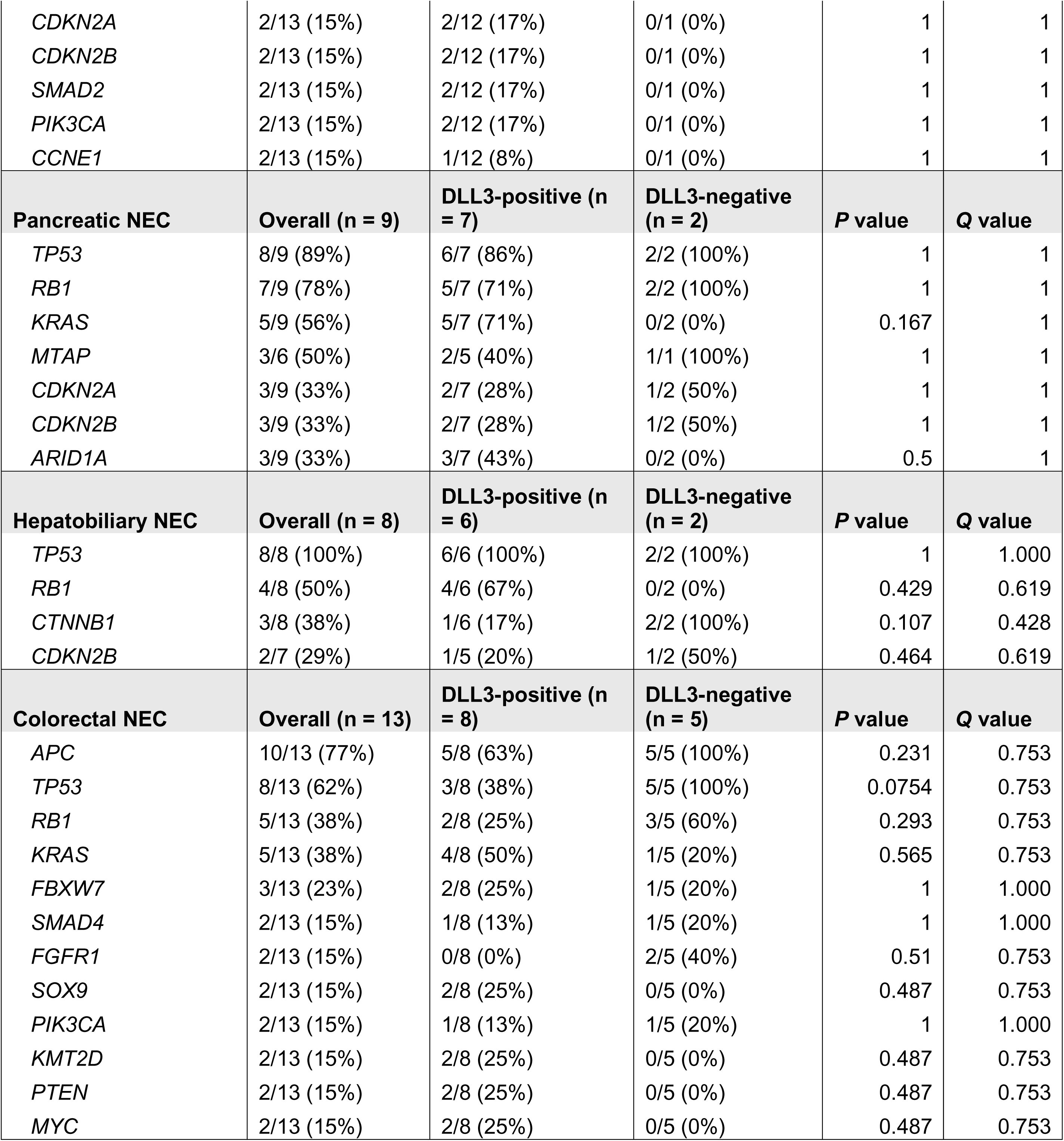
Genomics of poorly differentiated gastroenteropancreatic neuroendocrine carcinomas (GEP NECs) by DLL3 immunohistochemistry (IHC) status. Tumors with available DLL3 IHC and MSK-IMPACT next generation sequencing results are included. For the overall GEP NEC cohort, only genes with at least 5% oncogenic or likely oncogenic alterations in the cohort overall are displayed. For individual GEP NEC subtypes, only genes with at least two samples with oncogenic alterations within the cohort are displayed. TMB = tumor mutational burden. IQR = interquartile range. MSI = microsatellite instability. MSI = microsatellite instability-high. MSS = microsatellite stable. NA = not available. *P* values correspond to Fisher’s exact test by gene between groups. *Q* values were calculated to account for multiple hypothesis testing.

**Supplementary Table 3:**
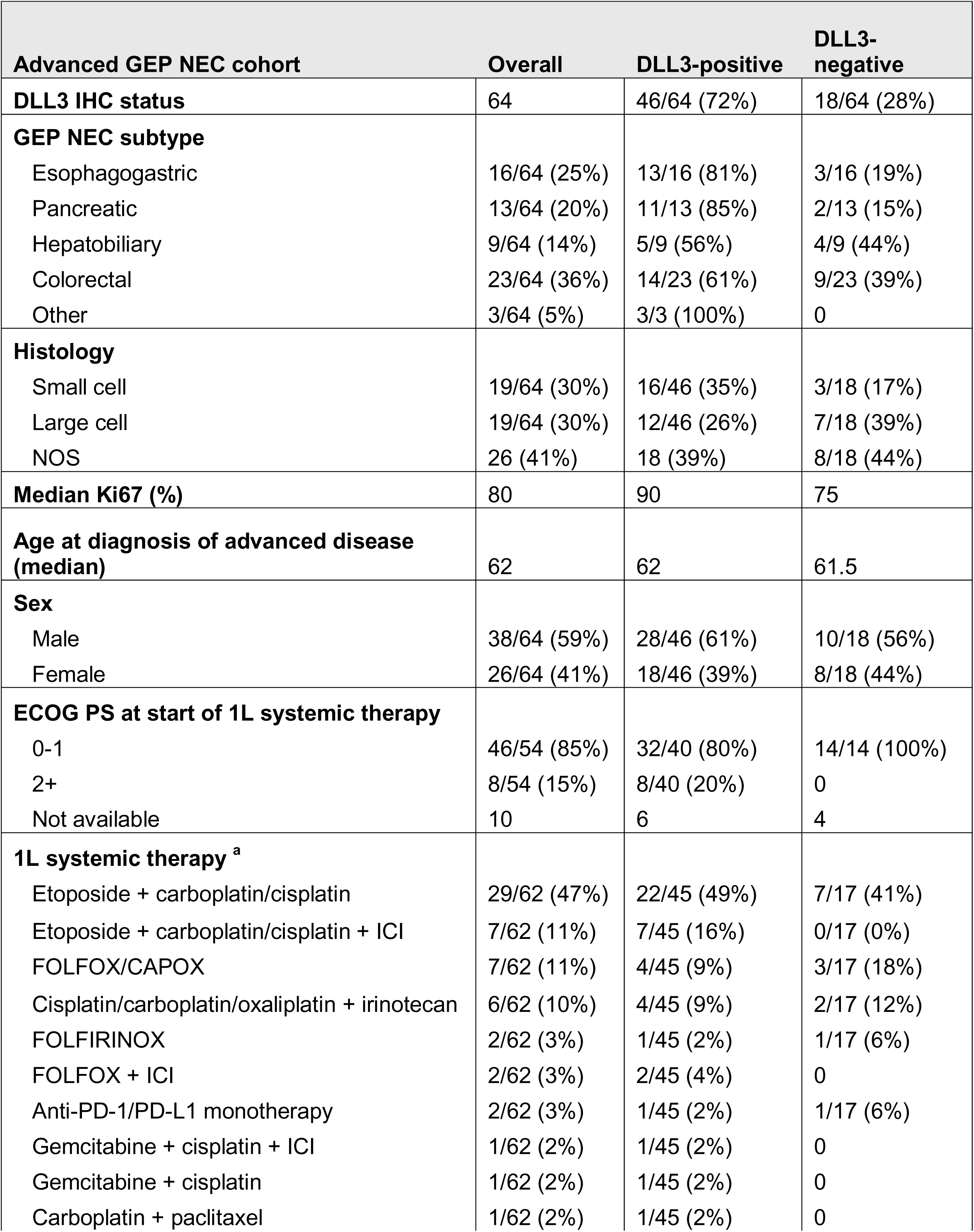
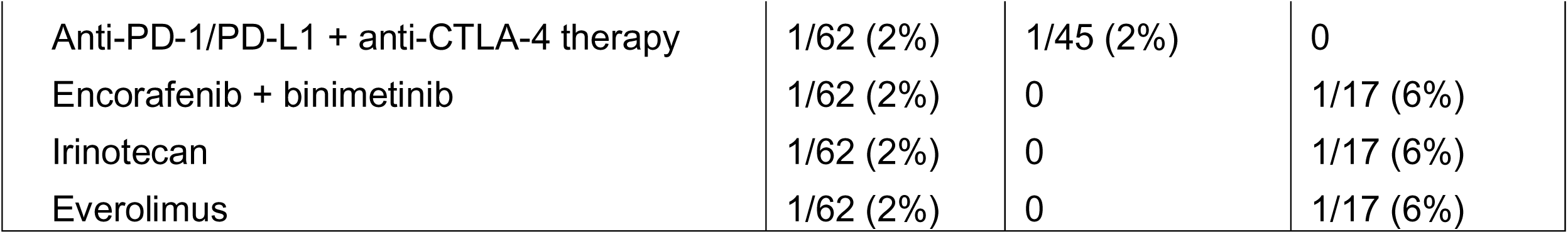
Clinical and pathologic characteristics of advanced poorly differentiated gastroenteropancreatic neuroendocrine carcinomas (GEP NEC) cohort. A total of 64 patients with advanced GEP NECs profiled by DLL3 immunohistochemistry (IHC) were included in the analysis. NOS = not otherwise specified. ECOG PS = Eastern Cooperative Oncology Group Performance Status. 1L = first-line. ICI = immune checkpoint inhibitor. Note: two patients did not receive systemic therapy for advanced disease (DLL3-positive = 1, DLL3-negative = 1).

**Supplementary Table 4:** Genomics of grade 3 pancreatic NETs (G3 PanNETs) by DLL3 immunohistochemistry (IHC) status. Tumors with available DLL3 IHC and MSK-IMPACT next generation sequencing results are included. Only genes with at least 5% oncogenic alterations in the cohort overall are displayed. *P* values correspond to Fisher’s exact test by gene between groups. *Q* values were calculated to account for multiple hypothesis testing.

| **G3 PanNET** | **Overall (n = 38)** | **DLL3-positive (n = 18)** | **DLL3-negative (n = 20)** | **P-value** | **Q-value** |
| --- | --- | --- | --- | --- | --- |
| *CDKN2A* | 18/38 (47%) | 10/18 (56%) | 8/20 (40%) | 0.516 | 1 |
| *MEN1* | 16/38 (42%) | 7/18 (39%) | 9/20 (45%) | 0.752 | 1 |
| *CDKN2B* | 13/38 (34%) | 7/18 (39%) | 6/20 (30%) | 0.734 | 1 |
| *DAXX* | 13/38 (34%) | 4/18 (22%) | 7/20 (35%) | 0.485 | 1 |
| *TSC2* | 8/38 (21%) | 3/18 (17%) | 5/20 (25%) | 0.697 | 1 |
| *MTAP* | 4/21 (19%) | 1/9 (11%) | 3/12 (25%) | 0.603 | 1 |
| *TP53* | 6/38 (16%) | 3/18 (17%) | 3/20 (15%) | 1.000 | 1 |
| *SETD2* | 4/38 (11%) | 2/18 (11%) | 2/20 (10%) | 1.000 | 1 |
| *ATRX* | 3/38 (8%) | 1/18 (6%) | 2/20 (10%) | 1.000 | 1 |
| *PTEN* | 3/38 (8%) | 0/18 (0%) | 3/20 (15%) | 0.232 | 1 |
| *KRAS* | 3/38 (8%) | 1/18 (6%) | 2/20 (10%) | 1.000 | 1 |

**Supplementary Table 5:** Clinical and pathologic characteristics of advanced pancreatic neuroendocrine tumor (PanNET) cohort. A total of 66 patients with advanced PanNETs profiled by DLL3 immunohistochemistry (IHC) were included in the analysis. G = grade. ECOG PS = Eastern Cooperative Oncology Group Performance Status. 1L = first-line. SSA = somatostatin analog. ICI = immune checkpoint inhibitor. 2L = second-line. Notes: (a) one patient (DLL3-negative) did not receive systemic therapy for advanced disease, and treatment information was not available for one patient (DLL3-negative). (b) 13 patients did not receive 2L systemic therapy for advanced disease. (c) 8 patients did not receive systemic therapy after diagnosis of G3 advanced disease. (d) 20 patients did not receive 2L systemic therapy for G3 advanced disease.

| **Advanced PanNET cohort** | **Overall** | **DLL3-positive** | **DLL3-negative** |
| --- | --- | --- | --- |
| **DLL3 IHC status** | 66 | 18/66 (27%) | 48/66 (73%) |
| **Tumor grade at DLL3 IHC evaluation** |  |  |  |
| G1 | 8/66 (12%) | 0/8 (0%) | 8/8 (100%) |
| G2 | 13/66 (20%) | 1/13 (8%) | 12/13 (92%) |
| G3 | 45/66 (68%) | 17/45 (38%) | 28/45 (62%) |
| **Median Ki67 at DLL3 IHC evaluation (%)** | 25 | 50 | 23 |
| **Age at diagnosis of advanced disease (median)** | 59 | 59 | 58.5 |
| **Sex** |  |  |  |
| Male | 41/66 (62%) | 7/18 (39%) | 34/48 (71%) |
| Female | 25/66 (38%) | 11/18 (61%) | 14/48 (29%) |
| **Hormone secreting/functional disease** |  |  |  |
| Functional | 12/64 (19%) | 5/18 (28%) | 7/46 (15%) |
| Non-functional | 52/64 (81%) | 13/18 (72%) | 39/46 (85%) |
| Not known | 2 | 0 | 2 |
| **ECOG PS at start of 1L systemic therapy for advanced disease** |  |  |  |
| 0-1 | 34/36 (94%) | 9/10 (90%) | 25/26 (96%) |
| 2+ | 2/36 (6%) | 1/10 (10%) | 1/26 (4%) |
| Not available | 30 | 8 | 22 |
| **Tumor grade at start of 1L systemic therapy for advanced disease <sup>a</sup>** |  |  |  |
| G1 | 20/65 (31%) | 3/18 (17%) | 17/47 (36%) |
| G2 | 27/65 (42%) | 7/18 (39%) | 20/47 (43%) |
| G3 | 18/65 (28%) | 8/18 (44%) | 10/47 (21%) |
| **1L systemic therapy for advanced disease (any grade) <sup>a</sup>** |  |  |  |
| SSA monotherapy | 38/64 (59%) | 11/18 (61%) | 27/46 (59%) |
| Capecitabine + temozolomide | 15/64 (23%) | 4/18 (22%) | 11/46 (24%) |
| Etoposide + carboplatin/cisplatin +/- SSA | 4 (6%) | 0 | 4/46 (9%) |
| Everolimus +/- SSA | 4 (6%) | 2/18 (11%) | 2/46 (4%) |
| Etoposide + carboplatin/cisplatin + ICI | 2/64 (3%) | 1/18 (6%) | 1/46 (2%) |
| FOLFOX | 1/64 (2%) | 0 | 1/46 (2%) |
| **2L systemic therapy for advanced disease (any grade) <sup>b</sup>** |  |  |  |
| Capecitabine + temozolomide | 16/53 (30%) | 5/18 (28%) | 11/35 (31%) |
| <sup>177</sup>Lu-DOTATATE | 15/53 (28%) | 2/18 (11%) | 13/35 (37%) |
| SSA monotherapy | 5/53 (9%) | 2/18 (11%) | 3/35 (9%) |
| Everolimus | 4/53 (8%) | 2/18 (11%) | 2/35 (6%) |
| Sunitinib | 3/53 (6%) | 3/18 (17%) | 0 |
| FOLFOX/CAPOX | 2/53 (4%) | 1/18 (6%) | 1/35 (3%) |
| Dacarbazine | 2/53 (4%) | 1/18 (6%) | 1/35 (3%) |
| Everolimus + CDK4/6 inhibitor | 1/53 (2%) | 1/18 (6%) | 0 |
| FOLFIRI | 1/53 (2%) | 0 | 1/35 (3%) |
| Gemcitabine + oxaliplatin | 1/53 (2%) | 1/18 (6%) | 0 |
| FOLFIRINOX | 1/53 (2%) | 0 | 1/35 (3%) |
| MK-0646 | 1/53 (2%) | 0 | 1/35 (3%) |
| Temozolomide | 1/53 (2%) | 0 | 1/35 (3%) |
| **1L systemic therapy for advanced G3 disease (G3 PanNET only) <sup>c</sup>** |  |  |  |
| <sup>177</sup>Lu-DOTATATE | 11/38 (29%) | 5/15 (33%) | 6/23 (26%) |
| Capecitabine + temozolomide | 9/38 (24%) | 4/15 (27%) | 5/23 (22%) |
| FOLFOX/CAPOX | 7/38 (18%) | 3/15 (20%) | 4/23 (17%) |
| SSA monotherapy | 6/38 (16%) | 2/15 (13%) | 4/23 (17%) |
| Etoposide + carboplatin/cisplatin + ICI | 2/38 (5%) | 1/15 (7%) | 1/23 (4%) |
| Etoposide + carboplatin/cisplatin | 1/38 (3%) | 0 | 1/23 (4%) |
| Gemcitabine + oxaliplatin | 1/38 (3%) | 0 | 1/23 (4%) |
| Cabozantinib | 1/38 (3%) | 0 | 1/23 (4%) |
| **2L systemic therapy for advanced G3 disease (G3 PanNET only) <sup>d</sup>** |  |  |  |
| FOLFOX/CAPOX | 8/26 (31%) | 3/11 (27%) | 5/15 (33%) |
| <sup>177</sup>Lu-DOTATATE | 6/26 (23%) | 1/11 (9%) | 5/15 (33%) |
| Capecitabine + temozolomide | 4 (15%) | 3/11 (27%) | 1/15 (7%) |
| SSA monotherapy | 3/26 (12%) | 2/11 (18%) | 1/15 (7%) |
| Etoposide + carboplatin/cisplatin | 1/26 (4%) | 1/11 (9%) | 0 |
| Everolimus | 1/26 (4%) | 0 | 1/15 (7%) |
| FOLFIRI | 1/26 (4%) | 0 | 1/15 (7%) |
| Gemcitabine + oxaliplatin | 1/26 (4%) | 1/11 (9%) | 0 |
| FOLFIRINOX | 1/26 (4%) | 0 | 1/15 (7%) |

**Supplementary Table 6:** Demographics and clinical characteristics of patients in whom DLL3 immunoPET-CT imaging was completed. NEC = poorly differentiated neuroendocrine carcinoma, G3 = grade 3, NET = well differentiated neuroendocrine tumor, CTCAE = Common Terminology Criteria for Adverse Events.

|  |  |
| --- | --- |
| **Median age at enrollment (range), years** | 55.5 (37-76) |
| **Sex** |  |
| Male | 4 (67%) |
| Female | 2 (33%) |
| **Race** |  |
| White | 4 (67%) |
| Asian | 1 (17%) |
| Native Hawaiian/Pacific Islander | 1 (17%) |
| **Diagnosis** |  |
| Pancreatic NEC | 1 (17%) |
| Gallbladder NEC | 1 (17%) |
| G3 pancreatic NET | 4 (67%) |
| **Imaging timepoints of DLL3 immunoPET-CT (days post-injection)** |  |
| Day 3 | 3 (50%) |
| Day 4 | 2 (33%) |
| Day 5 | 1 (17%) |
| **Adverse events (CTCAE grade)** |  |
| None (grade 0) | 6 (100%) |

