## Supplementary Figures 1-6 for "Delta-like ligand 3 expression and functional imaging in gastroenteropancreatic neuroendocrine neoplasms"

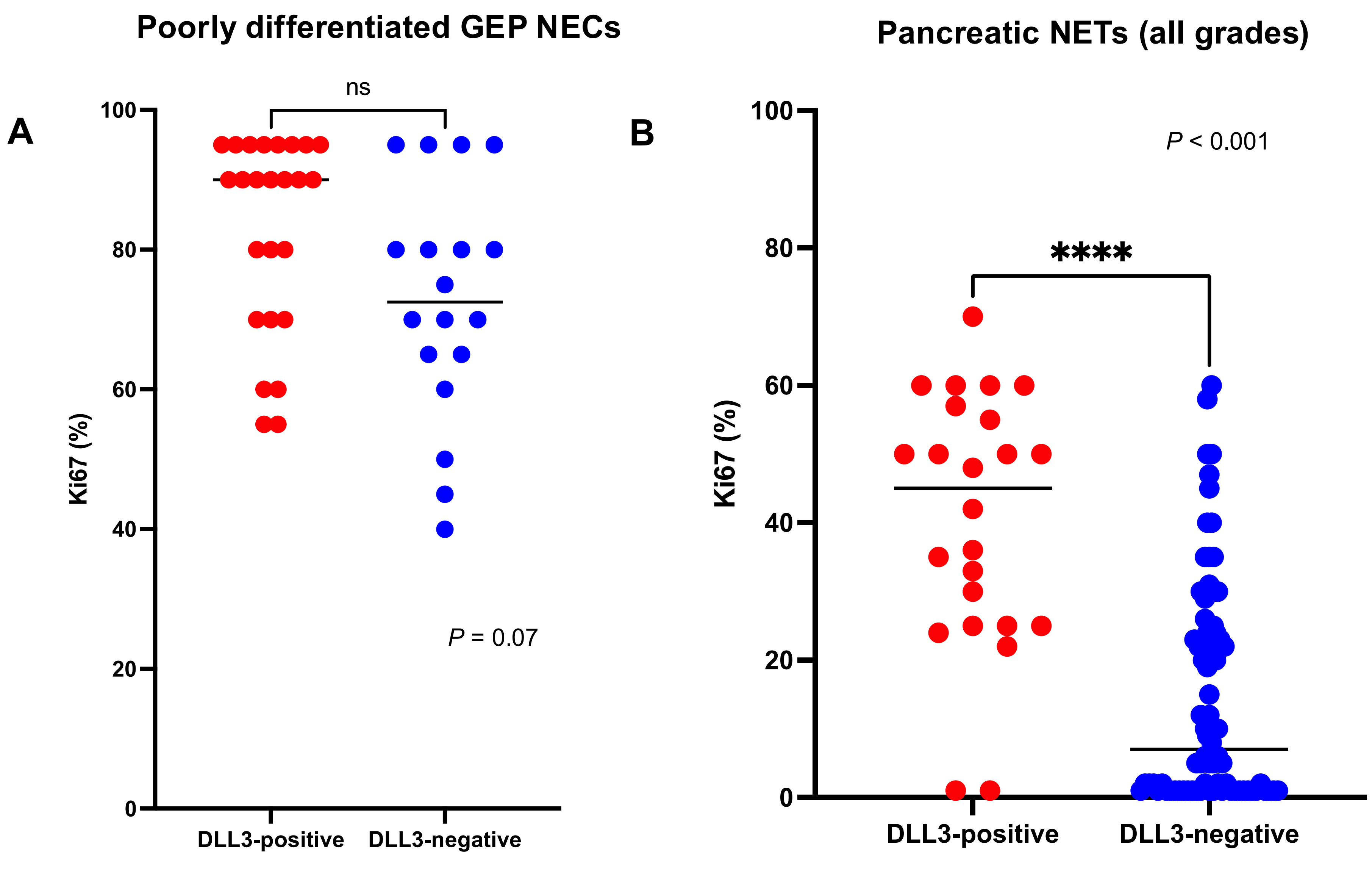


### Supplementary Figure 1

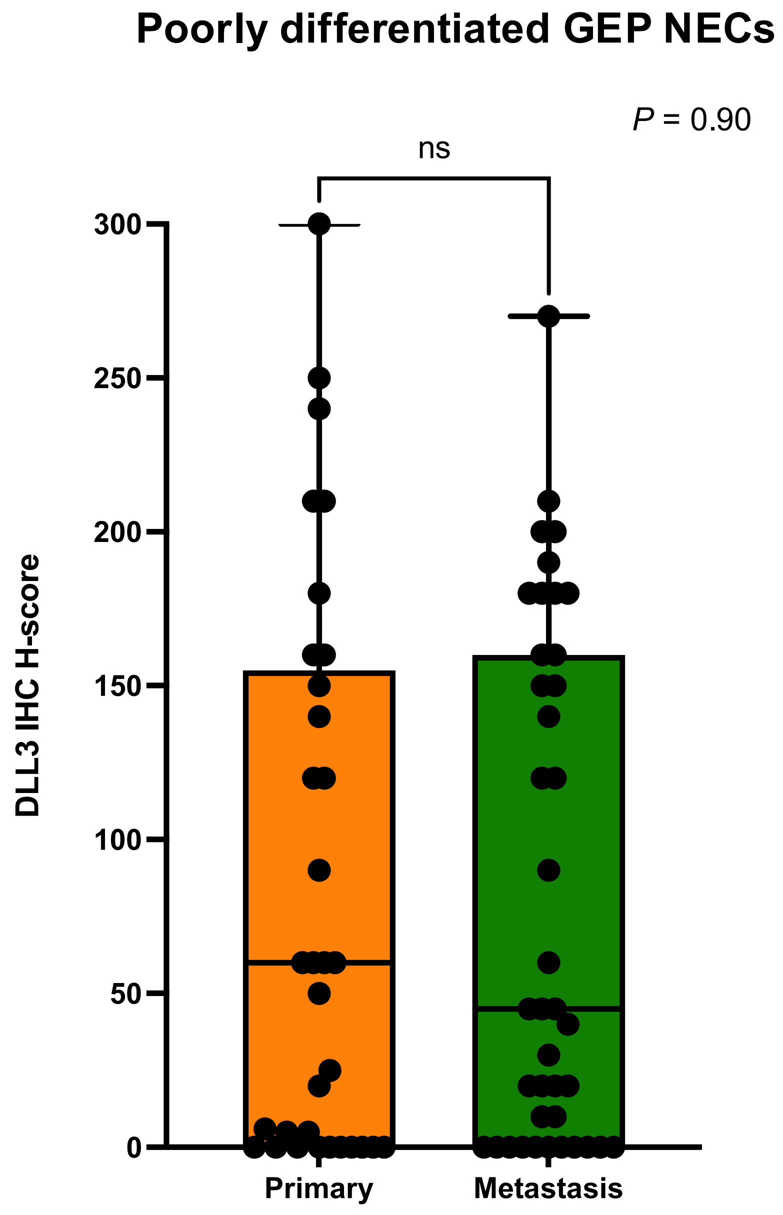


### Supplementary Figure 2

#### Poorly differentiated GEP NECs

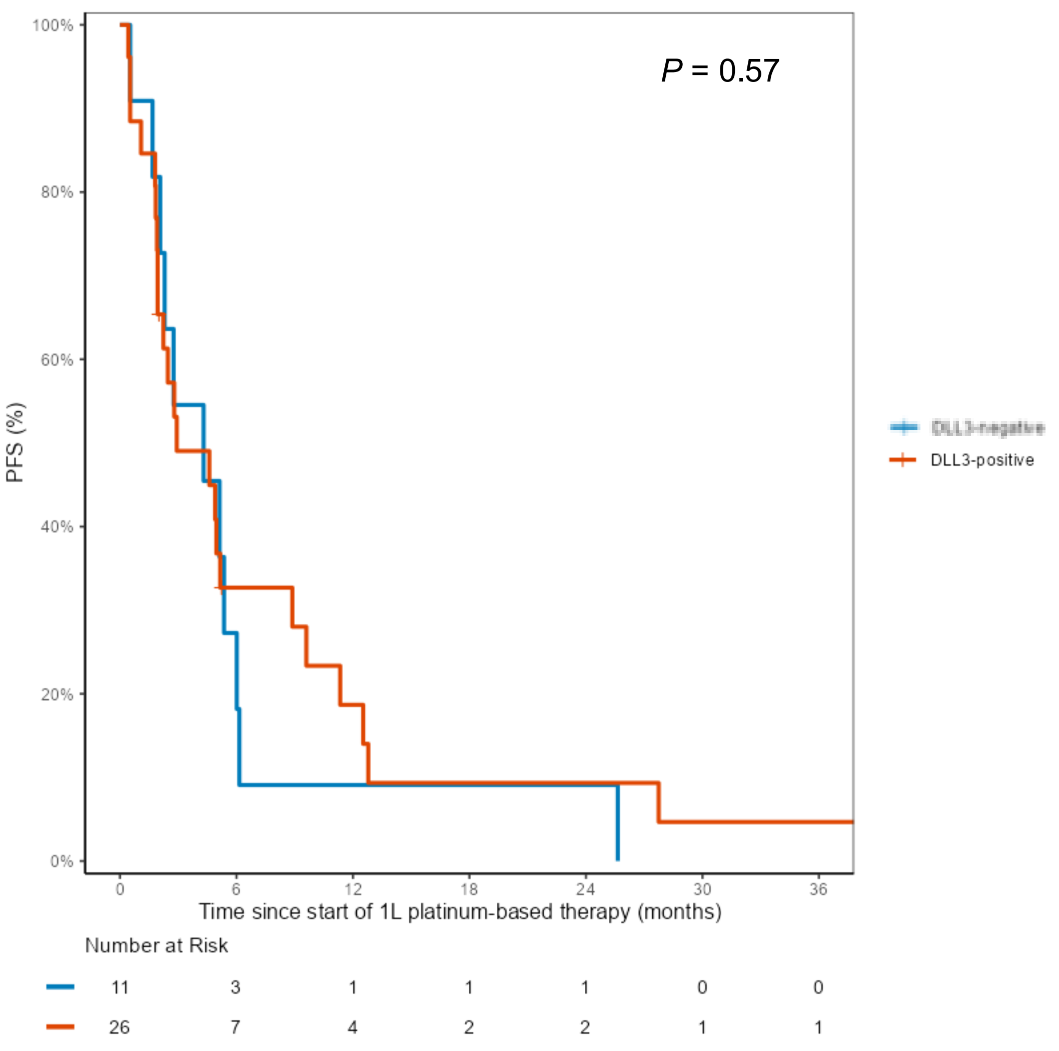


### Supplementary Figure 3

**Pancreatic NETs**

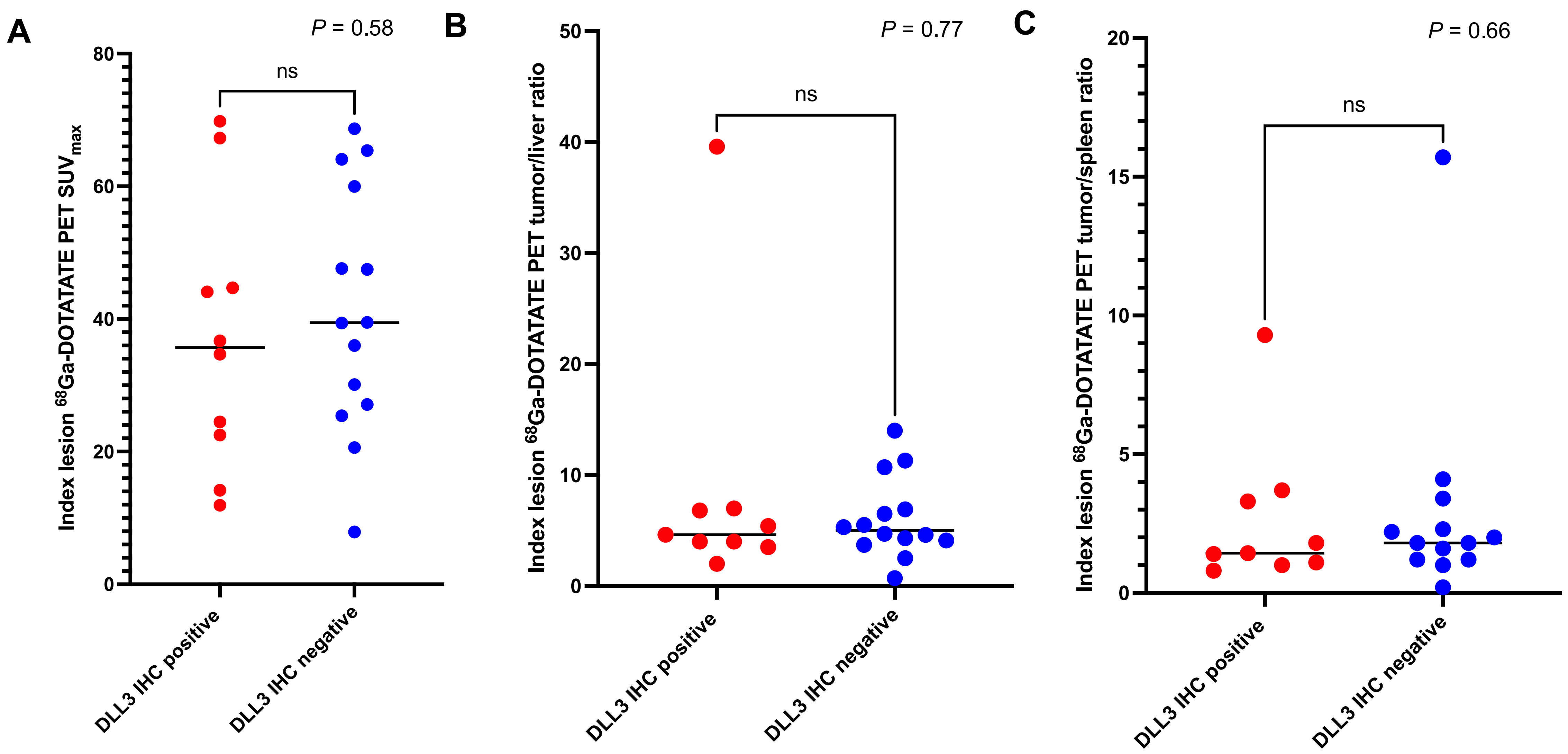


### Supplementary Figure 4

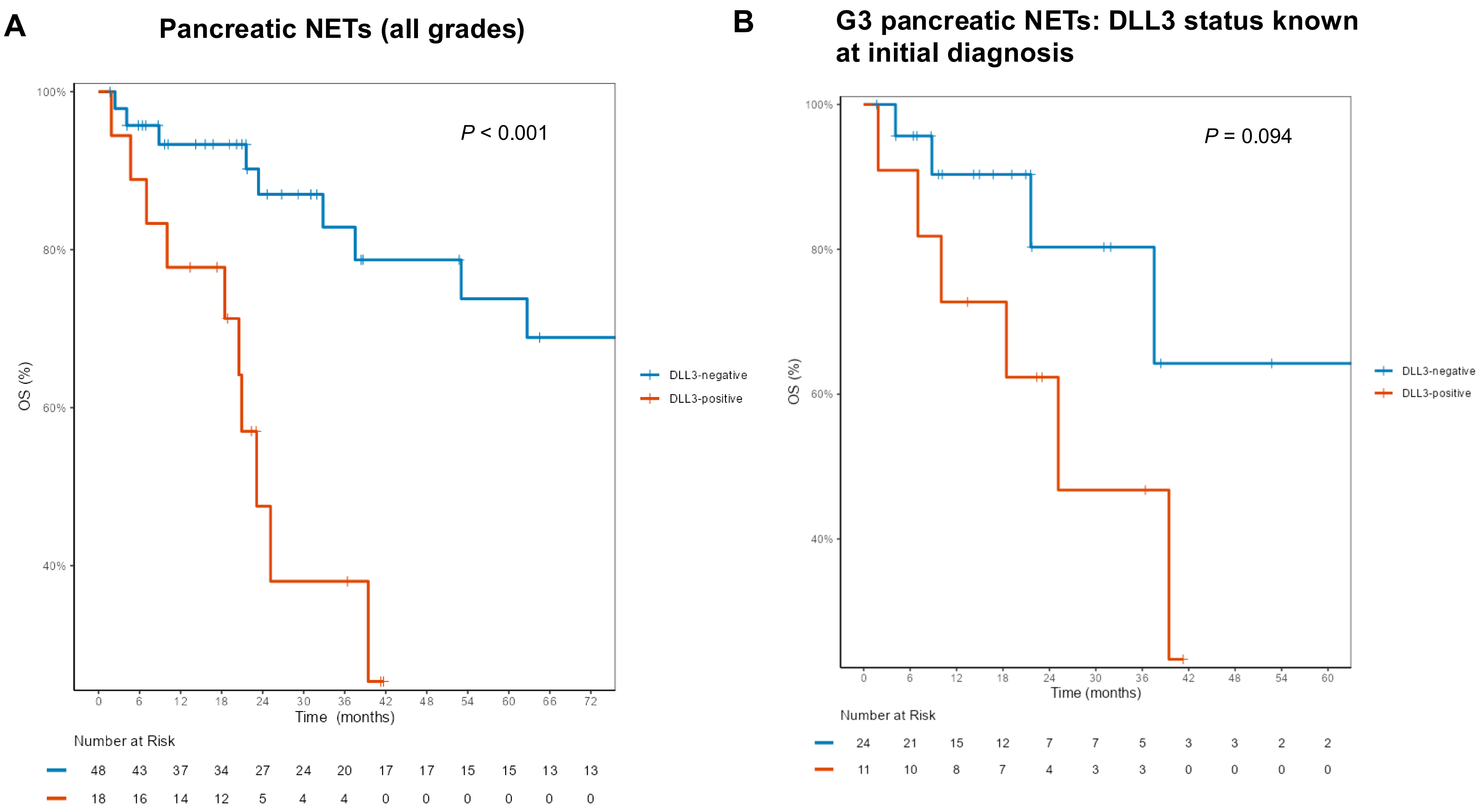


### Supplementary Figure 5

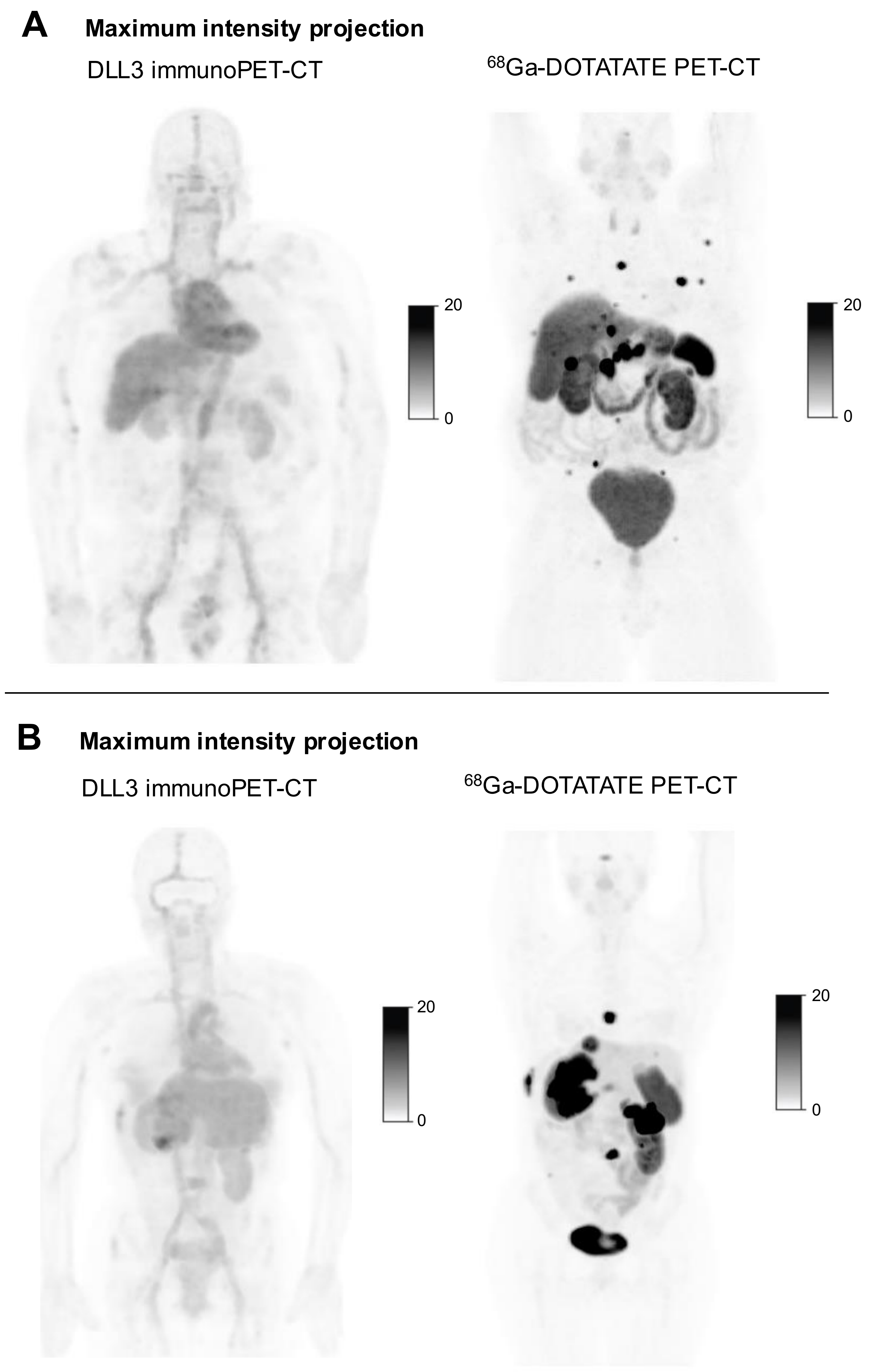


### Supplementary Figure 6
